# Measuring the impact of COVID-19 vaccination and immunity waning: a modelling study for Portugal

**DOI:** 10.1101/2021.12.10.21267618

**Authors:** Constantino Caetano, Maria Luísa Morgado, Paula Patrício, Andreia Leite, Ausenda Machado, André Torres, João Freitas Pereira, Sónia Namorado, Ana Sottomayor, André Peralta, Baltazar Nunes

## Abstract

Vaccination strategies to control COVID-19 have been ongoing worldwide since the end of 2020. Understanding their possible effect is key to prevent future disease spread. Using a modelling approach, this study intends to measure the impact of the COVID-19 Portuguese vaccination strategy on the effective reproduction number and explore three scenarios for vaccine effectiveness waning. Namely, the no-immunity-loss, 1-year and 3-years of immunity duration scenarios. We adapted an age-structured SEIR deterministic model and used Portuguese hospitalisation data for the model calibration. Results show that, although the Portuguese vaccination plan had a substantial impact in reducing overall transmission, it might not be sufficient to control disease spread. A significant vaccination coverage of those above 5 years old, a vaccine effectiveness against disease of at least 80% and softer non-pharmaceutical interventions (NPIs), such as mask usage and social distancing, would be necessary to control disease spread in the worst scenario considered. The immunity duration scenario of 1-year displays a resurgence of COVID-19 hospitalisations by the end of 2021, the same is observed in 3-year scenario although with a lower magnitude. The no-immunity-loss scenario presents a low increase in hospitalisations. In both the 1-year and 3-year scenarios, a vaccination boost of those above 65 years old would result in a 53% and 38% peak reduction of non-ICU hospitalisations, respectively. These results suggest that NPIs should not be fully phased-out but instead be combined with a fast booster vaccination strategy to reduce healthcare burden.

## 1 Introduction

During the pre-COVID-19 vaccine period, the pandemic has tested the capacity of health systems to deal with a high influx of COVID-19 patients. In order to reduce disease transmission, most governments opted to implement NPIs, such as general lockdowns, the closure of schools and the mandatory use of masks, among others. Portugal had two major lockdowns during this period. One at the middle of March 2020 during the first SARS-CoV-2 epidemic wave, and another one observed in January 2021, during the third epidemic wave [1]. By this latter date, the public health system was severely stressed with high numbers of hospitalisations. This surge of cases coincided with the appearance of the first Alpha-variant cases. This variant was the most commonly identified in the Portuguese population, until the identification of the new variant of concern (VOC) Delta. This new VOC became dominant, reaching 89.1% of confirmed COVID-19 cases in Portugal by the end of June 2021 [2]. It has been tied to higher disease transmission [3] and severe disease [4].

With the development of COVID-19 vaccines, the focus has changed from the sole use of NPIs to a mixture of both strategies. These vaccines have shown promising effects and mass vaccination programmes are underway worldwide. While such programmes are promising, several modelling studies have shown that as vaccination is rolled out, proper vaccine allocation is necessary [5, 6, 7, 8, 9] and maintenance of NPIs is still required to prevent the increase of hospitalisations and deaths, particularly before completing full vaccination [10, 11, 12, 13, 14]. In Portugal, vaccination has been rolled out in two main phases: the first focused in healthcare professionals and vulnerable individuals (residents in nursing homes, individuals at higher risk of severe disease due to preexisting medical conditions and those aged ≥ 80 years old), and the second one for the general population, organised by age groups, from 79 years old downwards. By 26th September, 84% of the population had received the complete vaccination scheme [15]. Four vaccines have been used: Cominarty (pfizer) and Spikevax (moderna), both on a 2-dose scheme with a 28 day interval, Vaxzevria (astrazeneca), using a 2-dose scheme with an interval from 8 to 12 weeks, and COVID-19 Janssen, with a single dose. These vaccines have been shown to be highly effective in the real-world. Vaccine effectiveness was estimated to be 89.1% against infection in fully vaccinated individuals and 99% against death [16]. Other studies have also shown that these vaccines might not grant long-lasting protection. Vaccine induced protection has been shown to last at least 5-6 months [17, 18, 19] and older individuals and at risk groups had higher rates of immunity loss [17]. Possible loss of immunity has led the Portuguese government to create new boosting vaccine strategies, giving priority to individuals most at risk of infection and severe disease, including healthcare professionals and older adults aged 65 or more.

Mathematical modelling and scenario development are tools that allow the simulation of the possible healthcare service impact by considering the introduction of new changes to COVID-19 dynamics, namely the appearance of a new VOC, vaccination strategies and immunity loss. Hence, our objectives with this project are twofold. First to analysed the impact of the Portuguese vaccination campaign on the effective reproduction number (*Rt*) and secondly to develop epidemic scenarios of trajectories of COVID-19 intensive care unit cases (ICU) and non-ICU hospitalisations regarding potential waning of vaccine induced protection and infection induced immunity. The results and conclusions from this study will help to support public health policy making in order to avoid future COVID-19 related healthcare burden.

## 2 Methods

We have considered a simplified version of the age-structured SEIR model developed as part of our previous work [20]. Details of the model are described in Appendix A. Individuals start off as susceptible (S) and, upon contact with an infectious individual, have a probability to become infected. Upon infection an individual is considered infectious after a period of time given by the average of the latency period for SARS-CoV-2 [21]. After this period an individual can either become symptomatic or asymptomatic with a given probability. Asymptomatic individuals infect at a reduced rate [22]. All asymptomatic individuals are assumed to recover from the infection while symptomatic individuals can either recover or require hospital admission. Hospitalised individuals can either recover, require intensive care or die. Individuals in intensive care units can either die or recover. To assess the effect of vaccination we added a further branch to the model: susceptible and recovered individuals are vaccinated at rate (?) and move to an analogous compartment in the vaccine protected branch of the model. Here we considered that the vaccine is leaky [23], i.e., susceptible individuals have reduced probability of being infected upon an infectious contact (1 − *e*) and if infected, have a reduced chance of developing symptoms (1 − *f*). Vaccinated individuals are similar in all manners to non-vaccinated individuals with the exception of the parameters mentioned above. See Appendices A and B for more information on model description and parameters.

In the simulations presented, individuals were vaccinated per age-group attempting to capture the progression of the national COVID-19 vaccination programme. Individuals in each age-group were assumed to be vaccinated according to the information available of vaccine coverage, planned vaccination schedule and vaccines recommended for the respective age-group. In the model, we assumed that an individual is protected by the vaccine a period after completing the vaccination scheme, assuming the targeted coverage described in Table 1 [15]. Age-groups not corresponding to the ones used in the model were assigned a coverage which is proportional to their size. For younger age groups a mixture of planned vaccination dates and potential vaccines to be administered were used. Beginning of vaccination in each age-group started two weeks after the beginning of administration of the vaccine in the previous group and was terminated 28 days after, assuming that each age-group received both COVID-19 Janssen vaccine or Spikevax /Cominarty vaccines. Non-calibrated parameters were obtained in COVID-19 literature and are described in Table 2 and presented in Table 3, Table 4, Table 5, Table 6 and Table 7, in Appendix B.

The duration of infection induced immunity and vaccine induced protection is present in the model and is given by the *γ* and *γ*_*v*_ parameters. An individual who loses its vaccine induced protection has the same characteristics as a non-vaccinated individual. In what follows, we will assume that both the infection induced immunity and vaccine induced protection have the same duration.

We fitted the model to the data on hospitalisations both in non-ICU and ICU, by age-group, starting on the January 24th 2021 and ending on September 29th 2021. Changes in contacts due to the introduction and lifting of NPIs and overall changes in population behaviour were estimated using a similar method described elsewhere [20, 24]. Details of the fitting procedure can be found in Appendix B.

In order to assess the impact of the Portuguese vaccination strategy on disease spread, we determined the *Rt* using formula (27). We have considered different levels of transmission associated with the presence of the Delta variant: *R*_0_ = 2.5, 3.2, 4.0 and 5.0 [3]. Vaccine effectiveness was considered to vary from 50% to 100% by varying the *e* parameter in equation (18) and fixing the parameter *f* as 0.5, in order to obtain the desired range. We also explored the *Rt* calculation with and without NPIs in place, such as the use of mask and social distancing. To this end, we assumed a 47% reduction in effective contacts according to mask effectiveness studies [25]. In these calculations we assumed that 11% of the population had already been infected and thus removed from the susceptible group [26]. The remaining susceptible population is then divided into vaccinated and non-vaccinated according to the vaccine coverage plan presented in Table 1. We have also considered the vaccination of those between the ages of 5-11, which is not currently part of the vaccination plan. We assumed a final coverage of 80% in this group. Other parameters used are described in Table 2.

**Table 1:**
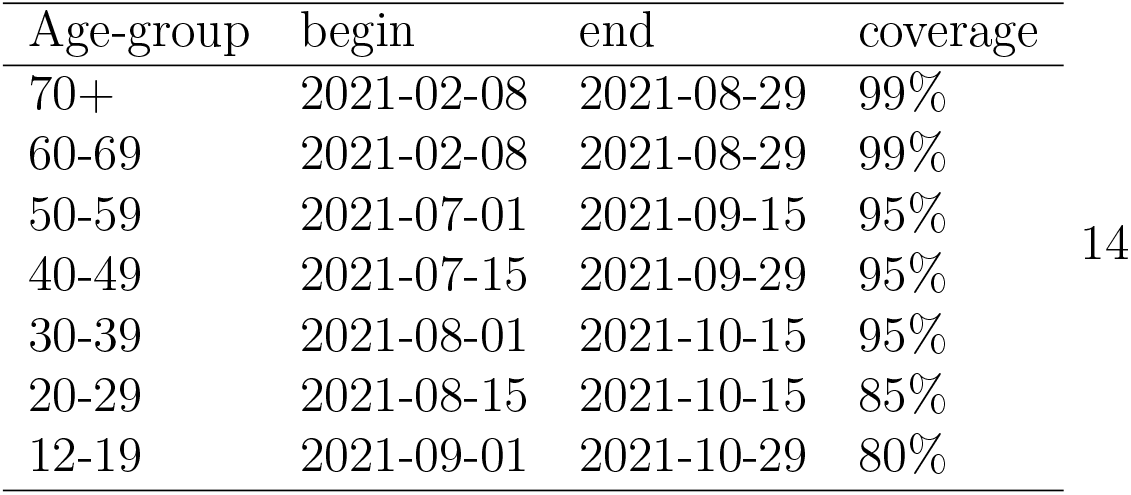
Portuguese vaccination roll-out plan. Beginning and end of the vaccination scheme by age-group and targeted final vaccination coverage [15].

Secondly, we developed scenarios of trajectories of COVID-19 hospitalisations, for three levels of vaccine induced protection waning. We explored a 1-year and a 3-years vaccine protection duration scenario and a no-loss-of-immunity scenario. Although little information is available about the duration of vaccine induced protection we chose these values according to recent studies that report at least 5-6 months of vaccine protection [17, 18, 19]. In each scenario we also explored the effect of giving a booster dose to those above 65 years of age. We assumed that this new vaccination scheme starts on October 11th 2021 and reaches 99% coverage within a month. We also included a 14-day delay to account for immune response.

The increased level of transmission due to the Delta variant was also included in the simulations. This was achieved by changing the value of the *β* parameter in formula (27), in order to be in line with the *R*_0_ estimated in COVID-19 literature [3] and fixing all other known parameters. For these simulations we assumed an *R*_0_ = 4 and that no NPIs were in place after September 30th 2021.

## 3 Results

In this section we present the results for the *Rt* impact values and the waning immunity scenarios.

### 3.1 Rt impact

Figure 1 depicts the *Rt* calculations for two scenarios with different levels of vaccine effectiveness, vaccination coverage and an increase of the reproduction number due to the circulation of the Delta variant, 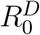. We can observe in both scenarios that administering the vaccine to more age-groups (y-axis) has a considerable impact in reducing the effective reproduction number. Moreover, this reduction is more noticeable when all population with 5 or more years is vaccinated.

**Figure 1:**
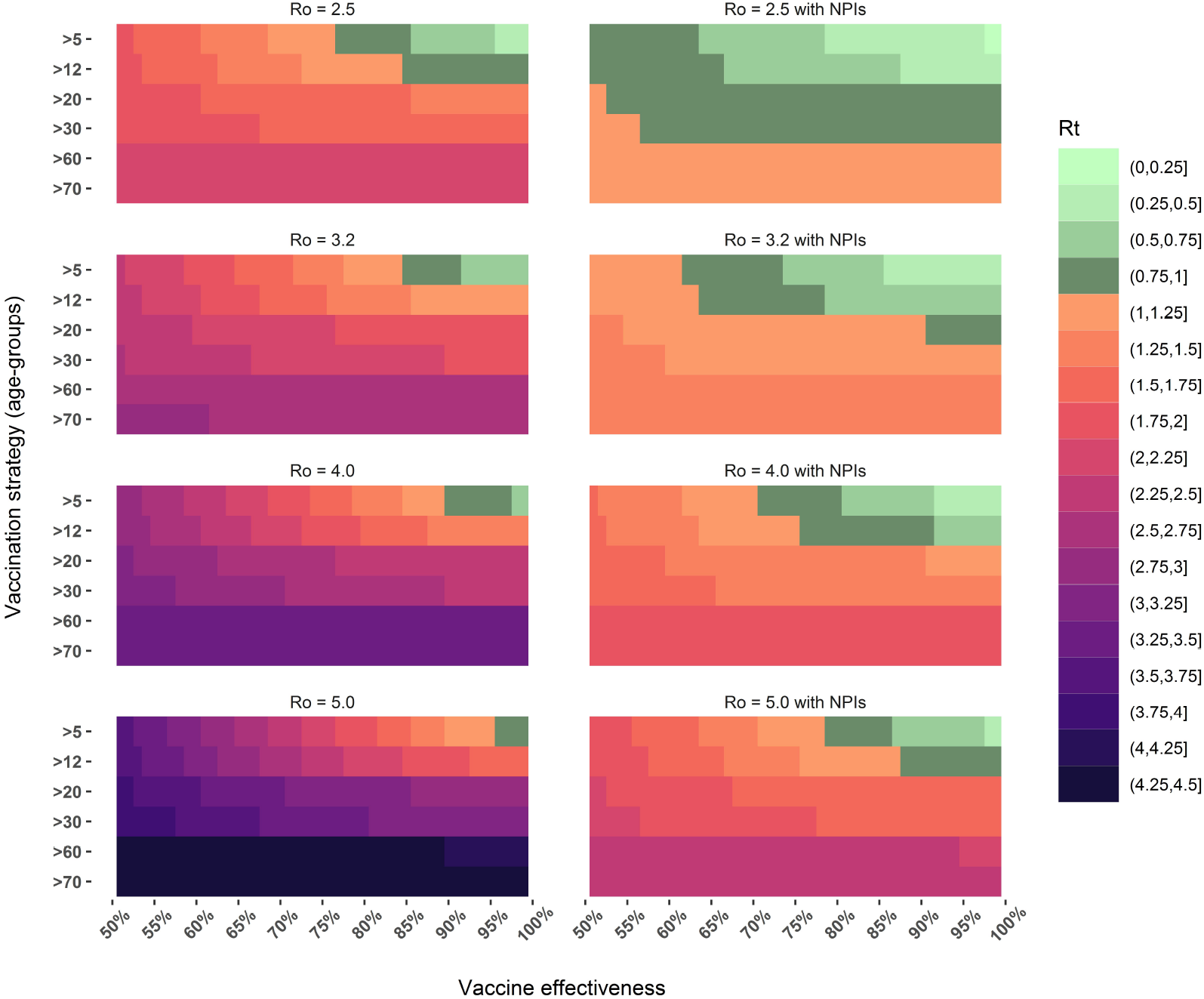
Effective reproduction number calculation as a function of vaccine coverage (by age-group), vaccine effectiveness and basic reproduction number. Two scenarios were considered: no NPIs in place (left) and overall effective contacts reduced by 47% (right) to take into account social distancing and mask usage [25]. The vaccination coverage was assigned to each age-group according to the final targeted coverage in the Portuguese vaccination plan, presented in Table 1.

Figure 1 describes the *Rt* in both scenarios considered, i.e. with and without NPIs in place. In the scenario with no NPIs, we can observe that values of *Rt* below 1 can only be achieved with the vaccination of at least the age-groups above 12 years old, a vaccine effectiveness above 85% and considering that the Delta variant has a similar transmission dynamics as the original strain 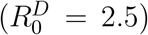 [27, 28]. The results for the scenario with NPIs in place show that an *Rt* below one can be achieved for all considered levels of transmission. For example, assuming the reproduction number of the Delta variant to be 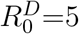, the effective reproduction number below 1 is achievable for vaccine effectiveness values above 80%, vaccination of those with 5 or more years of age and with NPIs that induce a 47% reduction in the effective number of contacts, as depicted in panel (b) of 1.

### 3.2 Wane simulations

Figure 2 presents the results for the vaccine protection duration scenarios and booster shots. The 1-year immunity duration scenario displays a new wave of non-ICU hospitalisations at the end of 2021, that can be mitigated by 53% at its peak if a booster shot is administered to those above 65 years of age. The 3-year immunity scenario displays a new wave of hospitalisations with a much smaller magnitude then the previous mentioned scenario but similar to the January 2021 wave. In this scenario the administration of the booster shot to those above 65 years old can reduce the peak non-ICU hospitalisation cases by 38%. The no-immunity-loss scenario also displays a new wave of hospitalisations by the end of 2021, although with a much smaller magnitude than the previous scenarios. It does not exceed 1000 non-ICU occupied beds at its peak. Similar results are obtained for ICU occupations as depicted in figure 3. Only in the no-immunity-loss scenario is expected not to exceed the maximum acceptable occupancy of ICU beds (255) by the end of 2021 [29]. More detailed results by age-group of the fitting procedure and simulation scenarios are presented in Appendix B.

**Figure 2:**
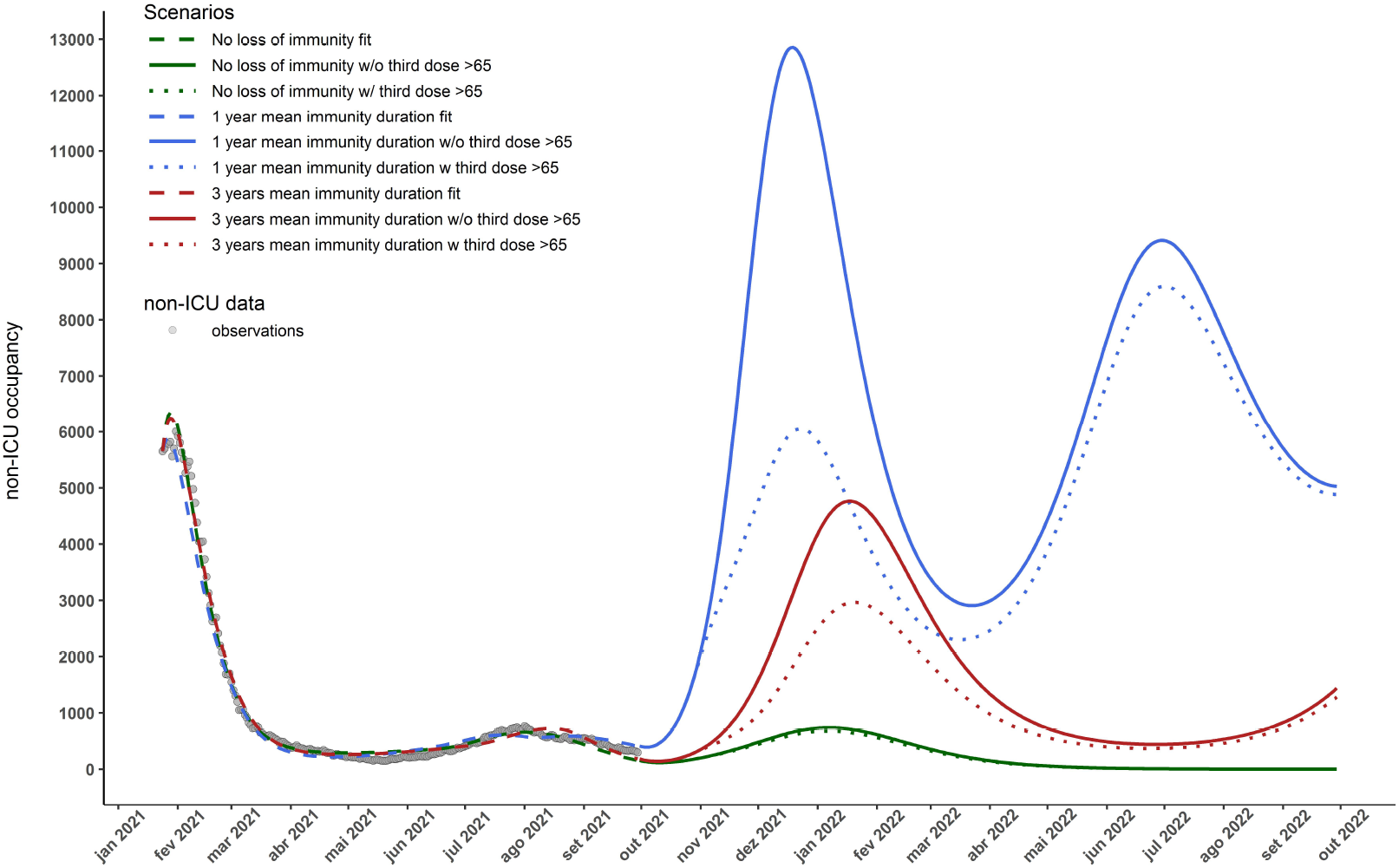
Number of COVID-19 non-ICU hospitalised cases for the 3 considered scenarios: no loss of vaccine induced protection (green), vaccine effectiveness duration of 1 and 3 years (blue and red respectively). Dashed lines depict the model calibration for each immunity loss scenario, the solid lines represent the trajectory of the total number of non-ICU hospitalisation cases in each scenario. The dotted lines represent the trajectories where a third dose of the vaccine was administrated to those above 65 years old and assuming that this group achieves 99% coverage with this new dose within one month of October 11th 2021. It is also assumed that no NPIs are in place after September 30th 2021 and that vaccine/infection granted immunity lasts an equal amount of time and is homogeneous among age-groups.

**Figure 3:**
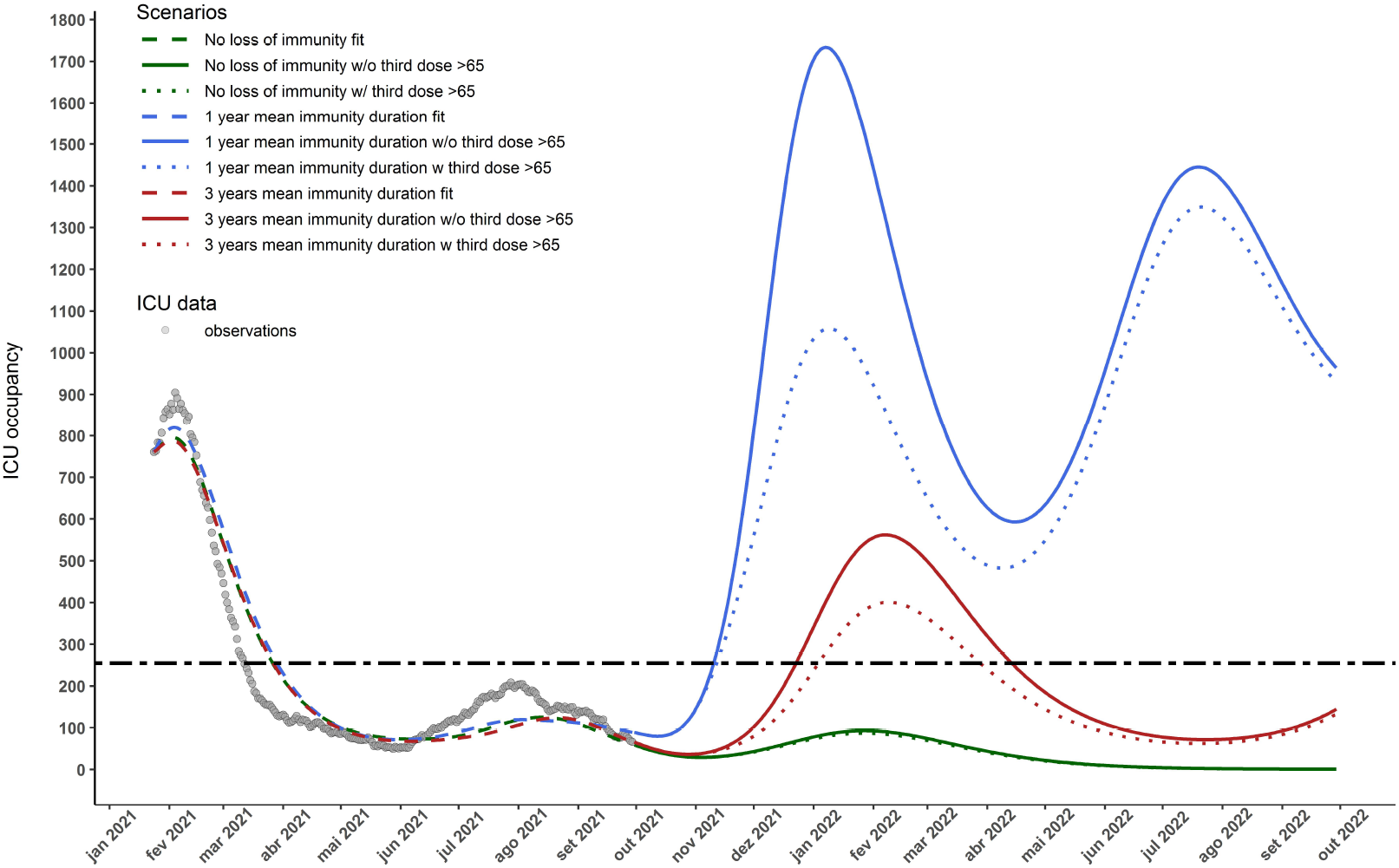
Number of COVID-19 ICU hospitalised cases for the 3 considered scenarios: no loss of vaccine granted protection (green), vaccine effectiveness duration of 1 and 3 years (blue and red respectively). Dashed lines depict the model calibration for each immunity loss scenario, the solid lines represent the trajectory of the total number of hospitalisation cases in each scenario. The dotted lines represent the trajectories where a third dose of the vaccine was administrated to those above 65 years old and assuming that this group achieves 99% coverage with this new dose within one month of October 11th. It is also assumed that no NPIs are in place after September 30th 2021 and that vaccine/infection granted immunity lasts an equal amount of time and is homogeneous among age-groups. The horizontal dashed line depicts the ICU capacity of COVID-19 hospitalisations in Portugal.

## 4 Discussion

The proposed model attempted to capture the dynamics of COVID-19 spread during the period from January 24th to September 30th 2021. It takes into account the heterogeneous nature of the Portuguese vaccination plan, the lifting and implementation of NPIs and the characteristics of disease complications in each age-group.

The *Rt* calculations performed enabled the analysis of possible outcomes of disease spread according to the vaccine effectiveness, vaccine allocation and increased transmission associated with the new Delta variant. By considering a different array of possible values for each of these parameters we were able to explore the *Rt* uncertainty and investigate possible outcomes of disease transmission. These results suggest that the herd-immunity threshold [30] might be higher than expected according to early assumptions due to the presence of new and more transmissible variants and herd-immunity might also not be attainable since vaccines do not grant immunity to infection. Recent COVID-19 reports in Portugal highlight that the *Rt* was estimated as 1.11 for the period between November 29th and December 3rd, vaccination coverage is above 85% in all eligible age-groups and early vaccine effectiveness studies estimate an effectiveness greater than 80% against COVID-19 related hospitalisation and mortality and are also effective against SARS-CoV-2 infection with rates of more then 53%. Hence, considering the present setting, our results suggest that solely vaccinating the targeted age-groups might not be sufficient to bring the *Rt* bellow one and prevent epidemic disease spread, even considering high vaccination coverage rates. These results are also in accordance with other Portuguese modelling studies [10, 31] and also international studies [11, 12, 13, 14] which also present epidemic spread in the presence of vaccination. Consequently the Portuguese government recently decided to vaccinate those between 5 to 11 years old, which according to our results would further reduce disease transmission and could even control disease spread in the worst hypothetical scenario considered (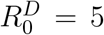 and no NPIs) if vaccine effectiveness is high (*>* 95%). Additionally under the hypothesis of vaccine immunity waning with time, one must consider that vaccine effectiveness changes throughout the pandemic, independently of vaccine coverage. This means that the impact of vaccine effectiveness and coverage on transmission can be transient in time. Being higher when effectiveness is high (after a booster dose campaign) and lower after more than 5-6 months of complete vaccination scheme. If epidemic control cannot be attained under these conditions, other measures should be adopted. The *Rt* impact results considering the presence of NPIs show that epidemic control might be feasible under the present setting if soft NPIs are maintained, such as the use of mask and social distancing. We can observe that an *Rt <* 1 can be achieved in the worst scenario 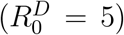 with those *>* 5 years old vaccinated and a vaccine effectiveness above 80%. Furthermore, we plan to update parameter values in these calculations as more detailed information becomes available regarding VOC transmission parameters.

The immunity-loss simulations proposed enabled us to quantify the future numbers of COVID-19 hospitalisations and ICU cases with regard to the uncertain nature of vaccine induced immunity waning and a new planned booster shot. These results show that a new wave of COVID-19 hospitalisations is expected by the end of 2021, and its magnitude is related to the duration of vaccine protection. We also showed that this wave can be heavily mitigated by administering a booster shot to those above 65 years of age (53% reduction at peak in non-ICU hospitalisations). The 1-year immunity loss scenario displays a possible wave of higher magnitude than past others in both non-ICU and ICU occupied beds, reaching 12857 non-ICU occupied beds at peak. This is a consequence of immunity loss of those that were vaccinated first, which included older and more at risk of severe disease individuals. In this scenario we can observe that administering a booster shot to those above 65 years old can substantially reduce the total number of hospitalisations by COVID-19. Note also that after this wave a new resurgence is expected by mid 2022 and in this wave the effect of the booster shot is dampened due to the immunity loss of those who received the booster dose. It is also important to state that we considered that loss of immunity after the booster dose is also 1 or 3 years, according to each scenario, this will be update as new data becomes available. The 3-year immunity duration scenario displays a resurgence of hospitalisations similar to the one observed during the beginning of 2021. In this case, giving a booster to those above 65 years of age also substantially reduces the number of hospitalised individuals (38%). The no-immunity-loss scenario depicts a small size wave of hospitalisations, and since vaccine grants long lasting immunity, the booster shot does not substantially impact the number of hospitalisations. Similar results have been reported by modelling studies that consider the presence of immunity waning, i.e., the presence of immunity loss might result in larger outbreaks, especially in the presence of a VOC with higher transmission [32, 33]. Another study suggests that if the rate of administration of booster shots is not high enough, it might lead to a resurgence of cases, that can be worse than previous waves [34]. It is important to note that our simulations assume a maximum potential of disease spread (*R*_0_ = 4), before vaccination and susceptibility and also that vaccine/infection induced immunity waning is age-group independent and no NPIs are introduced, even the use of face mask. This value was chosen according to estimated values of the reproduction numbers for the Delta variant [3]. A limitation of the analysis is that vaccinated individuals which lose their vaccine induced protection are assumed to have the same characteristics as non-vaccinated individuals within the same age-group. In a similar way, we assume that individuals who receive a booster shot are equally protected to those who have been vaccinated with a full scheme. Recent studies suggest that a third-dose booster shot increases vaccine protection to levels similar to receiving a complete scheme [35, 36] which is in line with the present assumptions.

Our results show that vaccine effectiveness, coverage, vaccination speed and vaccine induced protection waning play key roles in understanding disease spread and healthcare impact. Currently, vaccine allocation, effectiveness and coverage in Portugal has not been enough to control disease spread. Hence, NPIs should be maintained and combined with vaccination programmes that include more age-groups and booster doses. Vaccine effectiveness should be monitored in order to assess scenarios for the periodicity of booster shots. As more data becomes available we will extend the present model to study optimal booster dose schedules according to different scenarios of vaccine induced protection waning and infection induced immunity waning and the impact of new VOC.

## Data Availability

All data produced in the present study are available upon reasonable request to the authors.

## Appendix A Mathematical model

### Model description

A simplified version of our previous model for COVID-19 [20] was changed in order to accommodate vaccination and vaccine protection waning.

The homogeneous version of the model is then described through the following system of ODEs:

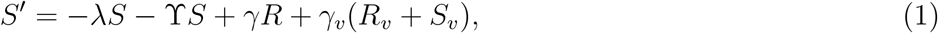

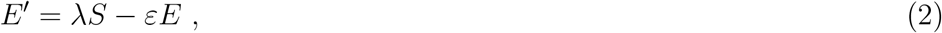

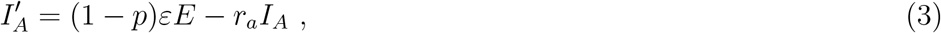

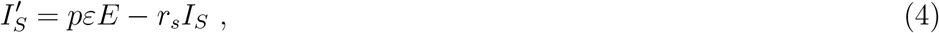

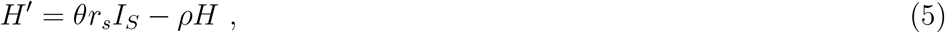

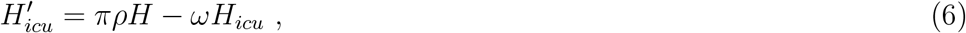

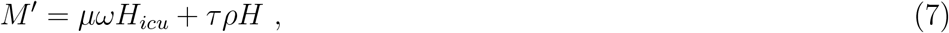

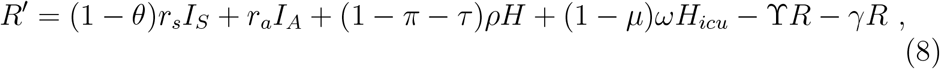

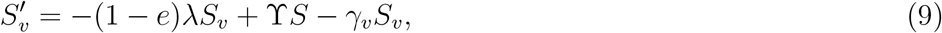

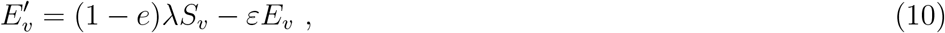

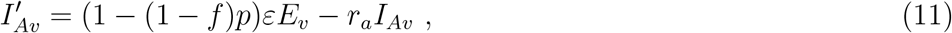

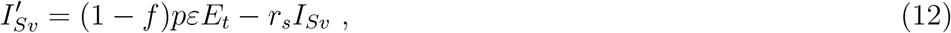

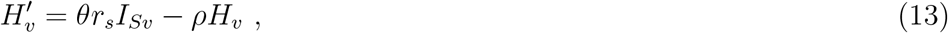

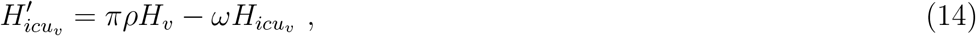

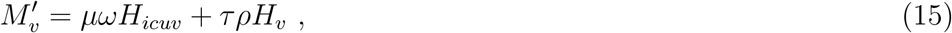

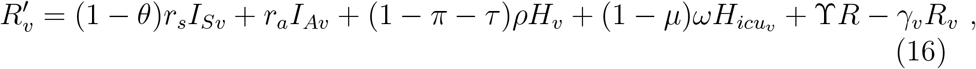

where *λ* = *β/N* (*α*_*S*_*C*_*S*_(*I*_*S*_ + *I*_*Sv*_) + *α*_*A*_*C*_*A*_(*I*_*A*_ + *I*_*Av*_)), *α*_*i*_ and *C*_*i*_, for *i* = *S, A*, are the transmissibility and contacts of symptomatic and asymptomatic infectious classes. Initial condition *S*(0), *E*(0), *I*_*A*_(0), *I*_*S*_(0), *H*(0), *H*_*icu*_(0), *M* (0), *R*(0), *S*_*v*_(0), *E*_*v*_(0), *I*_*Av*_(0), *I*_*Sv*_(0), *H*_*v*_(0), 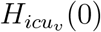, *M*_*v*_(0), *R*_*v*_(0) ≥ 0, summing to *N*.

A schematic diagram of the transmission model is represented in Figure 4. Model parameters description is presented in Table 2.

**Figure 4:**
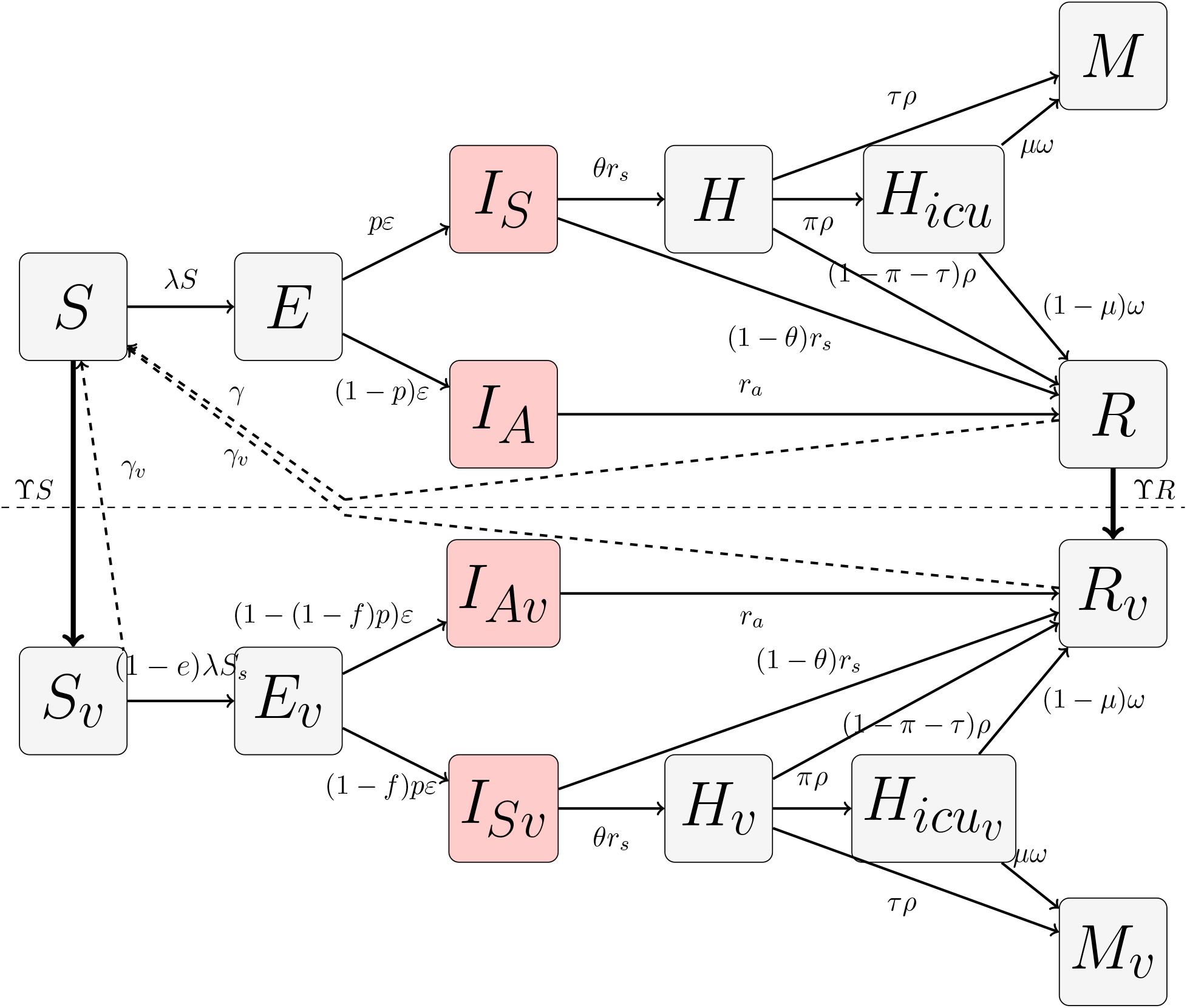
Schematic diagram of the homogeneous COVID-19 transmission model with vaccination. where *λ* = *β/N* (*α*_*S*_*C*_*S*_(*I*_*S*_ + *I*_*Sv*_) + *α*_*A*_*C*_*A*_(*I*_*A*_ + *I*_*Av*_), where *α*_*i*_ and *C*_*i*_, for *i* = *S, A*, are the relative transmissibility and contacts of infectious classes.

**Table 2:**
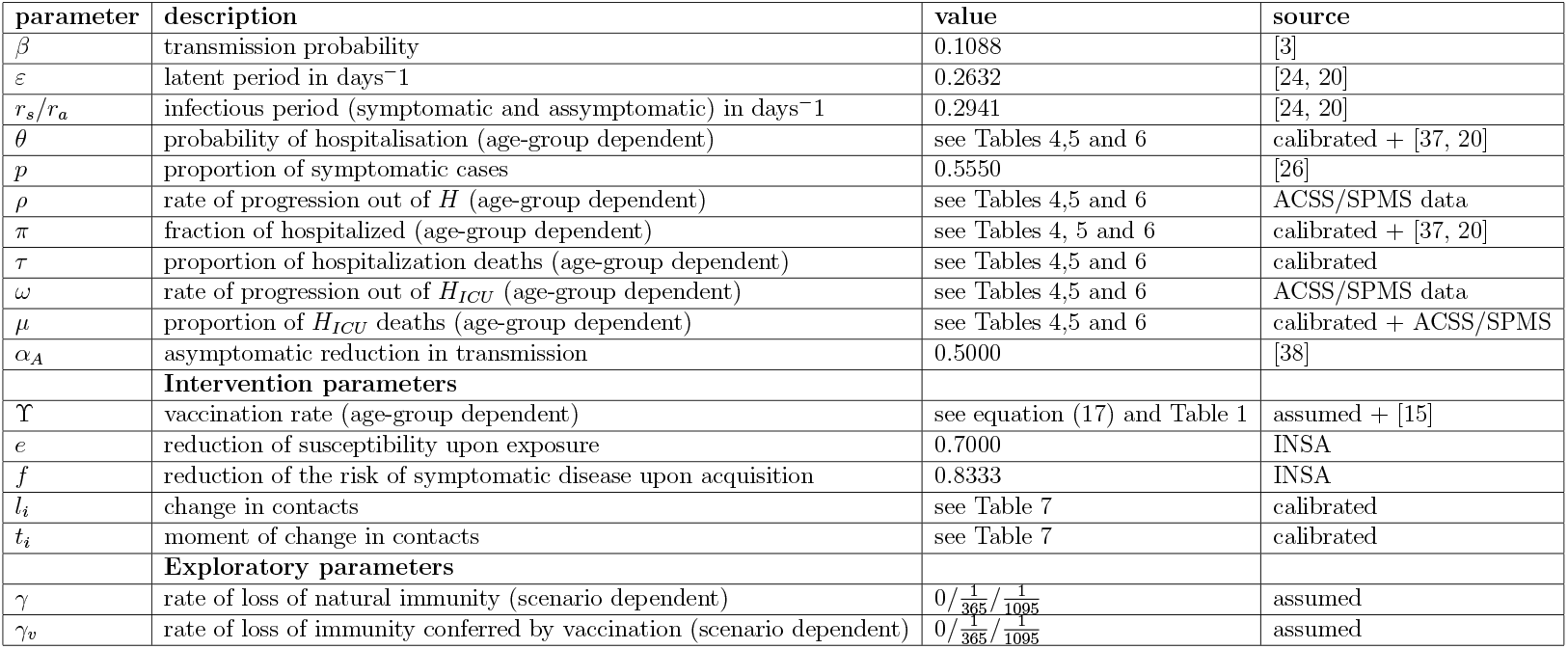
Description, value and source for the parameters used in the model’s simulations. INSA sourced parameters mean they were obtained from personal communication with the COVID-19 vaccine effectiveness study group at Instituto Nacional de Saúde Doutor Ricardo Jorge in Portugal. Parameters sourced as “calibrated + reference” mean that information about parameters found in literature were used to inform and calibrate the final value. ACSS/SPMS sourced parameters mean they were obtained from data analysis on hospitalisation data in Portugal.

| parameter | description | value | source |
| --- | --- | --- | --- |
| $\beta$ | transmission probability | 0.1088 | [3] |
| $\varepsilon$ | latent period in days<sup>-1</sup> | 0.2632 | [24, 20] |
| $r_s/r_a$ | infectious period (symptomatic and asymptomatic) in days<sup>-1</sup> | 0.2941 | [24, 20] |
| $\theta$ | probability of hospitalisation (age-group dependent) | see Tables 4,5 and 6 | calibrated + [37, 20] |
| $p$ | proportion of symptomatic cases | 0.5550 | [26] |
| $\rho$ | rate of progression out of $H$ (age-group dependent) | see Tables 4,5 and 6 | ACSS/SPMS data |
| $\pi$ | fraction of hospitalized (age-group dependent) | see Tables 4, 5 and 6 | calibrated + [37, 20] |
| $\tau$ | proportion of hospitalization deaths (age-group dependent) | see Tables 4,5 and 6 | calibrated |
| $\omega$ | rate of progression out of $H_{ICU}$ (age-group dependent) | see Tables 4,5 and 6 | ACSS/SPMS data |
| $\mu$ | proportion of $H_{ICU}$ deaths (age-group dependent) | see Tables 4,5 and 6 | calibrated + ACSS/SPMS |
| $\alpha_A$ | asymptomatic reduction in transmission | 0.5000 | [38] |
| **Intervention parameters** |  |  |  |
| $\Upsilon$ | vaccination rate (age-group dependent) | see equation (17) and Table 1 | assumed + [15] |
| $e$ | reduction of susceptibility upon exposure | 0.7000 | INSA |
| $f$ | reduction of the risk of symptomatic disease upon acquisition | 0.8333 | INSA |
| $l_i$ | change in contacts | see Table 7 | calibrated |
| $t_i$ | moment of change in contacts | see Table 7 | calibrated |
| **Exploratory parameters** |  |  |  |
| $\gamma$ | rate of loss of natural immunity (scenario dependent) | $0/\frac{1}{365}/\frac{1}{1095}$ | assumed |
| $\gamma_v$ | rate of loss of immunity conferred by vaccination (scenario dependent) | $0/\frac{1}{365}/\frac{1}{1095}$ | assumed |

For the heterogeneous version of the model we divide each compartment in *n* age classes. The force of infection is now:

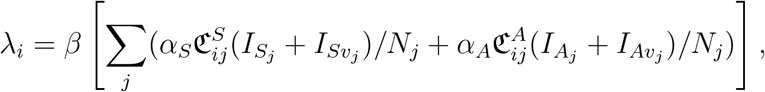

where *α*_*k*_ and 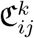, for *k* = *S, A*, are the relative transmission and contact matrices of the infectious classes, respectively. In our simulation study we consider 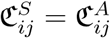.

### Vaccination effectiveness

For model simplicity we consider that only susceptible and recovered individuals are vaccinated. Vaccination of individuals is age dependent and according to the targeted vaccination program in Portugal (see Table 1). The vaccination rate is time dependent in a piecewise manner within each age-group and is given by the ϒ parameter. For each age-group ϒ_*i*_ is calculated as follows [31]:

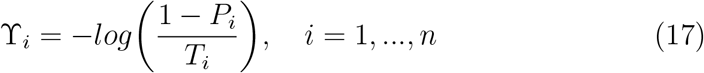

where *P*_*i*_ corresponds to the targeted final vaccination coverage in age-group *i* and *T*_*i*_ is the size of the time window of vaccination in age-group *i*.

As for vaccine effectiveness, imagine we are following a very small group for a short amount of time, we may assume that the force of infection is constant, *λ*^*^, so the risk of symptomatic disease upon exposure for a susceptible individual is

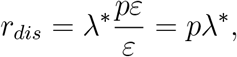

and for a vaccinated individual is

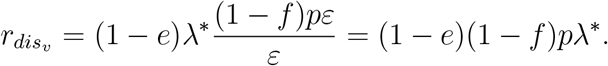

Hence the resulting reduction of the risk of symptomatic disease upon exposure is

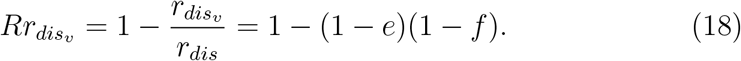

So the vaccine efficacy measured can result from different combinations of reductions of susceptibility upon exposure and of the risk of symptomatic disease upon acquisition.

### Effective reproduction numbers

The population is kept constant over the time, since it verifies

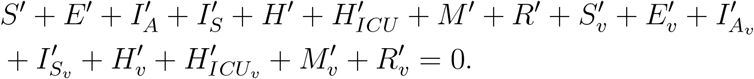

We take initial conditions *S*(0), *E*(0), *I*_*A*_(0), *I*_*S*_(0), *H*(0), *H*_*icu*_(0), *M* (0), *R*(0), *S*_*v*_(0), *E*_*v*_(0), *I*_*Av*_(0), *I*_*Sv*_(0), *H*_*v*_(0), 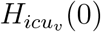, *M*_*v*_(0), *R*_*v*_(0) *≥* 0, summing to *N*. Assuming there is immunity waning, *γ*_*v*_ = *γ* ≠ 0, the disease free equilibrium has the form

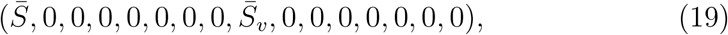

where 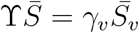. Assuming vaccination ? ≠ 0 we have

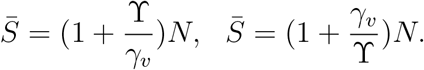

If we assume that vaccination is only administrated for a certain time interval [0, *T*] then 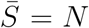 and 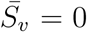. There is also an endemic solution for which 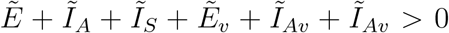. When considering that there is no loss of protection given by infection or disease, *γ*_*v*_ = *γ* = 0, all possible equilibria are disease free, of the form

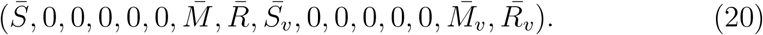

where 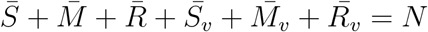.

To study the asymptotic behaviour of the model we linearize the system (1-16) around the disease free equilibria (19) or (20) following the *next generation approach* in [39]. The resulting linearized sub-system for the 6 infected classes is of the form

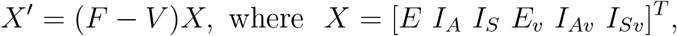

where the *F* matrix corresponds to new infections:

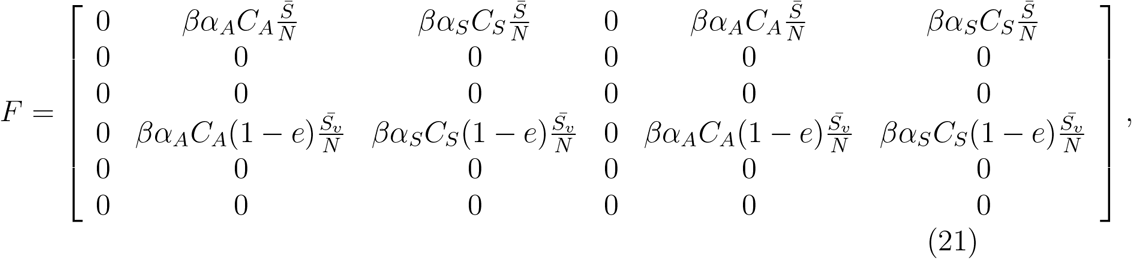

and *V* describes the other transitions between compartments

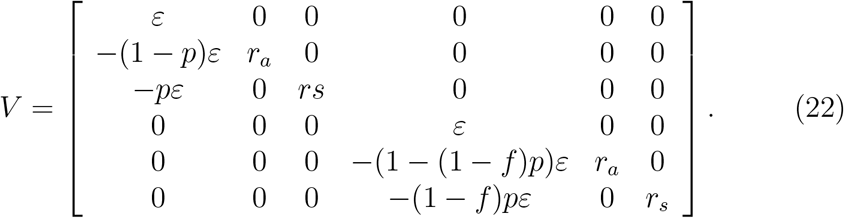

The next generation matrix is *FV* ^−1^ =

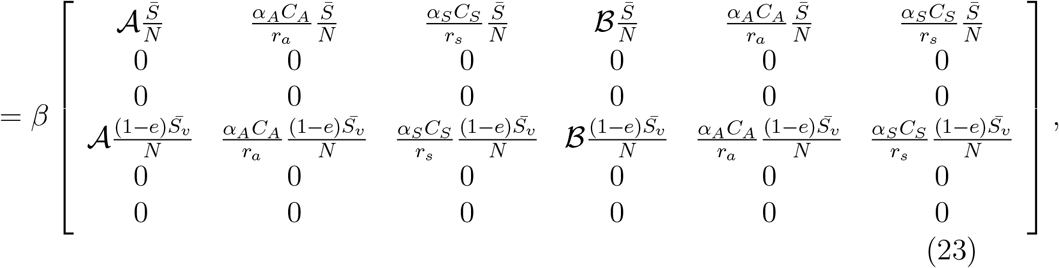

where 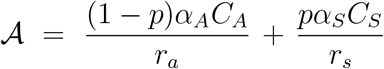 and 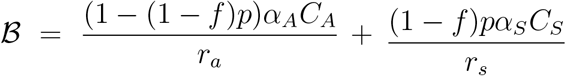. Hence, the effective reproduction number is the dominant eigenvalue of the next generation matrix

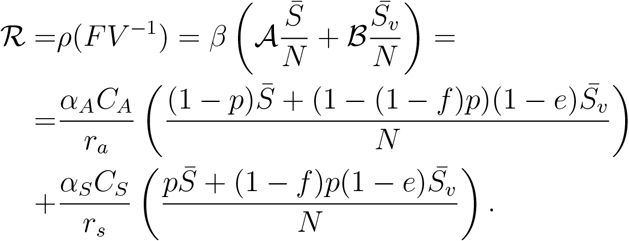

We can also define a time dependent effective reproduction number by replacing 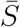 by *S*(*t*) and 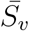 by *S*_*v*_(*t*) obtaining

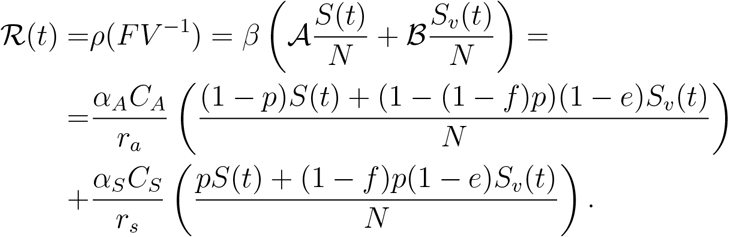

In the heterogeneous version of our COVID-19 vaccination model, we consider that each compartment is divided into *n* age classes to accommodate differences in contacts and disease related risks. Assuming waning of immunity, *γ*_*v*_ = *γ >* 0, there is a disease free equilibrium in each age class that is of the form

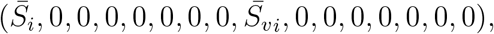

with 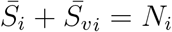and

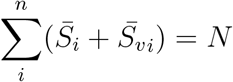

where *N*_*i*_ is the population size of age-group *i* and *N* is the total population.

When immunity is lifelong, *γ*_*v*_ = *γ* = 0, all possible equilibria are disease free of the form 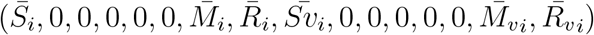, with 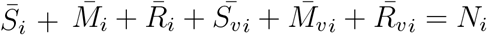 and

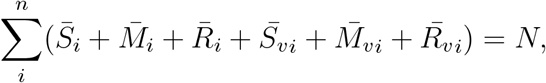

where *N*_*i*_ is the population size of age-group *i* and *N* is the total population.

As for the homogeneous model we linearize the system (1-8) following the next generation approach. The new infections matrix *F* is a *n* × *n* block matrix where each block is of the form:

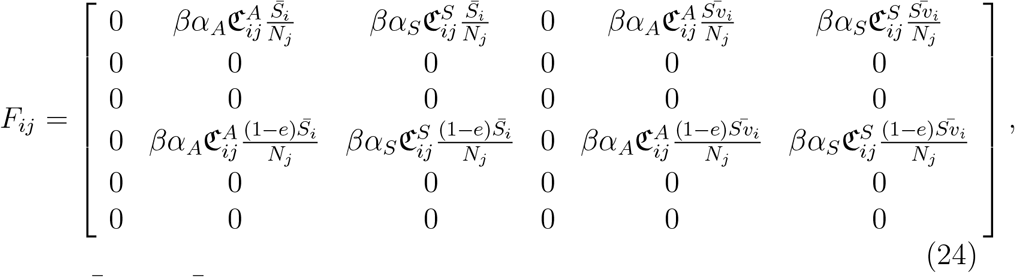

where 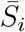 and 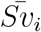 are the susceptible non vaccinated and vaccinated, respectively, at the disease free equilibrium considered of age-group *i, N*_*j*_ is the total population of age-group *j*, with *i, j* = 1, …, *n* and *n* is the number of age groups considered. *V* is a diagonal *n* block matrix where each block *i* is of the form:

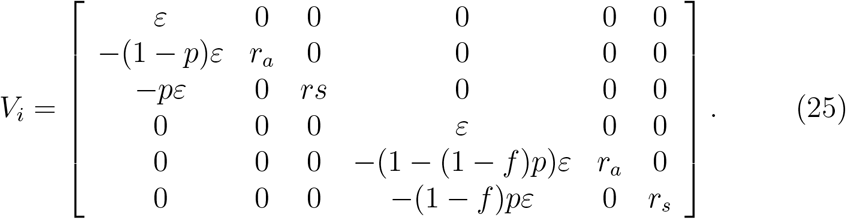

Hence, we define the effective reproduction number for the heterogeneous model as spectral radius of the next generation matrix:

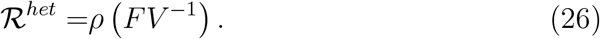

We can also define a time dependent effective reproduction number by replacing 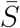 by *S*(*t*) and 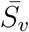 by *S*_*v*_(*t*) obtaining

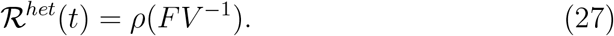

## Appendix B

### Data and Parameters

Data for the initial conditions on January 24th was obtained from different sources: Portuguese case data, deaths data and hospitalisation data. Contact data was obtained in [40]. Initial conditions are presented in Table 3. The model was calibrated on the number of non-ICU hospitalisations and individuals in ICU, this data was obtained from the ACSS/SPMS hospitalisation registry.

**Table 3:** Initial conditions for each age-group and non-vaccinated compartment for the starting simulation day of 2021-01-24. Initial conditions for all vaccinated compartments and age-groups is 0.

| Age group | S | E | IA | IS | H | ICU | M | R |
| --- | --- | --- | --- | --- | --- | --- | --- | --- |
| 0 - 4 | 386989.00 | 1144.00 | 811.00 | 1012.00 | 8.00 | 0.00 | 1.00 | 46237.00 |
| 5 - 9 | 402879.00 | 1631.00 | 1337.00 | 1668.00 | 9.00 | 0.00 | 0.00 | 48319.00 |
| 10 - 14 | 454523.00 | 2014.00 | 1472.00 | 1836.00 | 4.00 | 0.00 | 0.00 | 45091.00 |
| 15 - 19 | 490150.00 | 2436.00 | 1794.00 | 2238.00 | 5.00 | 0.00 | 2.00 | 48697.00 |
| 20 - 24 | 471175.00 | 2906.00 | 2030.00 | 2532.00 | 20.00 | 3.00 | 0.00 | 71778.00 |
| 25 - 29 | 468934.00 | 2764.00 | 2018.00 | 2517.00 | 20.00 | 2.00 | 8.00 | 71417.00 |
| 30 - 34 | 465720.00 | 2798.00 | 1987.00 | 2478.00 | 49.00 | 6.00 | 11.00 | 93545.00 |
| 35 - 39 | 552733.00 | 3263.00 | 2372.00 | 2958.00 | 59.00 | 7.00 | 13.00 | 111017.00 |
| 40 - 44 | 672346.00 | 3910.00 | 2769.00 | 3453.00 | 128.00 | 31.00 | 30.00 | 101557.00 |
| 45 - 49 | 676536.00 | 4130.00 | 2925.00 | 3648.00 | 129.00 | 32.00 | 63.00 | 102270.00 |
| 50 - 54 | 637053.00 | 3732.00 | 2649.00 | 3304.00 | 274.00 | 80.00 | 95.00 | 97991.00 |
| 55 - 59 | 633486.00 | 3535.00 | 2339.00 | 2917.00 | 272.00 | 79.00 | 184.00 | 97329.00 |
| 60 - 64 | 586943.00 | 3092.00 | 2004.00 | 2499.00 | 509.00 | 143.00 | 361.00 | 81211.00 |
| 65 - 69 | 541284.00 | 2295.00 | 1519.00 | 1895.00 | 468.00 | 132.00 | 570.00 | 74749.00 |
| 70 - 74 | 508977.00 | 1924.00 | 1170.00 | 1459.00 | 1226.00 | 81.00 | 844.00 | 33910.00 |
| 75+ | 1014532.00 | 6077.00 | 3433.00 | 4281.00 | 2473.00 | 164.00 | 8602.00 | 68359.00 |

In Table 2 we present a description, value and source for non-age dependent model parameters. Fixed and estimated age-group dependent parameters are presented in Table 4, Table 5 and Table 6.

Four periods were considered during the calibration window period (2021-01-24 to 2021-09-29). These were divided by moments when changes in contacts occurred. These were estimated as well as their associated contact changes. The results are depicted in Table 7.

### Likelihood function

The model was calibrated on hospitalisation data, both number of non-ICU hospitalisations and ICU hospitalisations. We consider that, on day *t*, for each age-group, the number of individuals in ICU and infirmary follow independent Poisson distribution with mean equal to *H*_*t*_ and 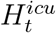, which correspond to the numerical solution for the number of hospitalised non-ICU cases (*H*+*H*_*v*_) and number of ICU hospitalisations 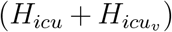 on day *t*. Hence model fitting was performed by minimising the following likelihood function on *θ*

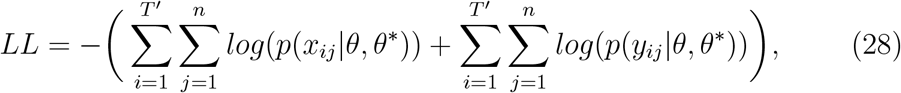

where *θ* is the vector of parameters we want to estimate, *θ*^*^ is the vector of known parameters, *x*_*ij*_ are the data on number of non-ICU cases of age *j* in day *t* = *i, y*_*ij*_ are the data on number of individuals in ICU of age *j* in day *t* = *i*. The function *p*(.) is the probability mass function of the Poisson distribution and *T* ^′^ is the window size in days of the calibration period. Fitting was performed via the differential evolution algorithm using the R package DEoptim [41].

#### 4.1 Fitting results

Figure 5 presents the model fitting for the total and age-group number of non-ICU and ICU cases for each scenario: no-immunity-loss, 1-year and 3-years immunity duration. The 1-year duration scenario presents the lowest negative log-likelihood (23782.15) followed by the 3-year immunity duration scenario (27811.8). The no-loss-immunity scenario presents the highest negative log-likelihood (31862.64), indicating a lesser fit to the data.

**Figure 5:**
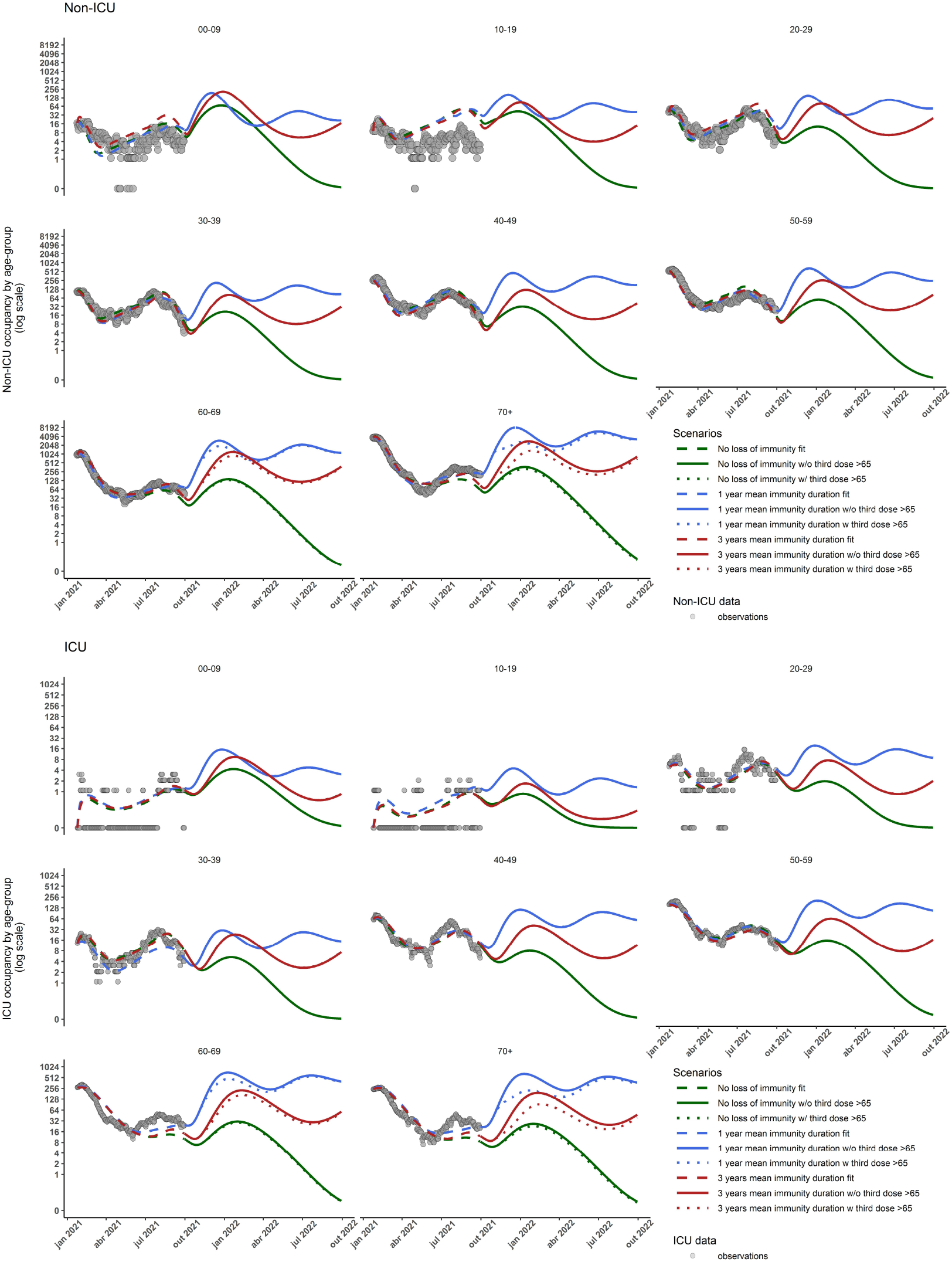
Results from the model fitting and booster dose simulations on non-ICU cases (top) and ICU cases (bottom) for each simulation scenario: no-immunity-loss scenario (*LL* = 31862.64), 1-year immunity duration scenario (*LL* = 23782.15) and 3-year immunity duration scenario (*LL* = 31862.64). All plots are in log2 scale.

Table 4, Table 5 and Table 6 show the estimated age-group dependent parameters for each scenario: (S1) no-immunity-loss scenario, (S2) 1-year immunity duration scenario and (S3) 3-year immunity duration scenario.

**Table 4:** Age-group dependent parameters, both estimated (highlighted) and fixed for the no-immunity-loss scenario.

| | Age groups | $\theta$ | $\pi$ | $\tau$ | $\mu$ | $\rho$ | $\omega$ |
| --- | --- | --- | --- | --- | --- | --- | --- |
| 1 | 0 - 4 | **0.01** | **0.04** | **0.00** | **0.10** | 0.17 | 0.03 |
| 2 | 5 - 9 | **0.01** | **0.00** | **0.00** | **0.00** | 0.17 | 0.03 |
| 3 | 10 - 14 | **0.01** | **0.02** | **0.00** | **0.00** | 0.12 | 0.07 |
| 4 | 15 - 19 | **0.01** | **0.00** | **0.00** | **0.00** | 0.12 | 0.07 |
| 5 | 20 - 24 | **0.01** | **0.03** | **0.01** | **0.00** | 0.14 | 0.05 |
| 6 | 25 - 29 | **0.01** | **0.07** | **0.00** | **0.00** | 0.14 | 0.05 |
| 7 | 30 - 34 | **0.01** | **0.07** | **0.00** | **0.00** | 0.12 | 0.06 |
| 8 | 35 - 39 | **0.01** | **0.18** | **0.00** | **0.11** | 0.12 | 0.06 |
| 9 | 40 - 44 | **0.01** | **0.11** | **0.02** | **0.07** | 0.10 | 0.05 |
| 10 | 45 - 49 | **0.01** | **0.15** | **0.03** | **0.04** | 0.10 | 0.05 |
| 11 | 50 - 54 | **0.03** | **0.16** | **0.01** | **0.16** | 0.09 | 0.04 |
| 12 | 55 - 59 | **0.03** | **0.13** | **0.06** | **0.14** | 0.09 | 0.04 |
| 13 | 60 - 64 | **0.15** | **0.12** | **0.07** | **0.24** | 0.08 | 0.04 |
| 14 | 65 - 69 | **0.15** | **0.13** | **0.08** | **0.24** | 0.08 | 0.04 |
| 15 | 70 - 74 | **0.28** | **0.08** | **0.18** | **0.34** | 0.07 | 0.04 |
| 16 | 75+ | **0.28** | **0.03** | **0.33** | **0.53** | 0.07 | 0.04 |

**Table 5:**
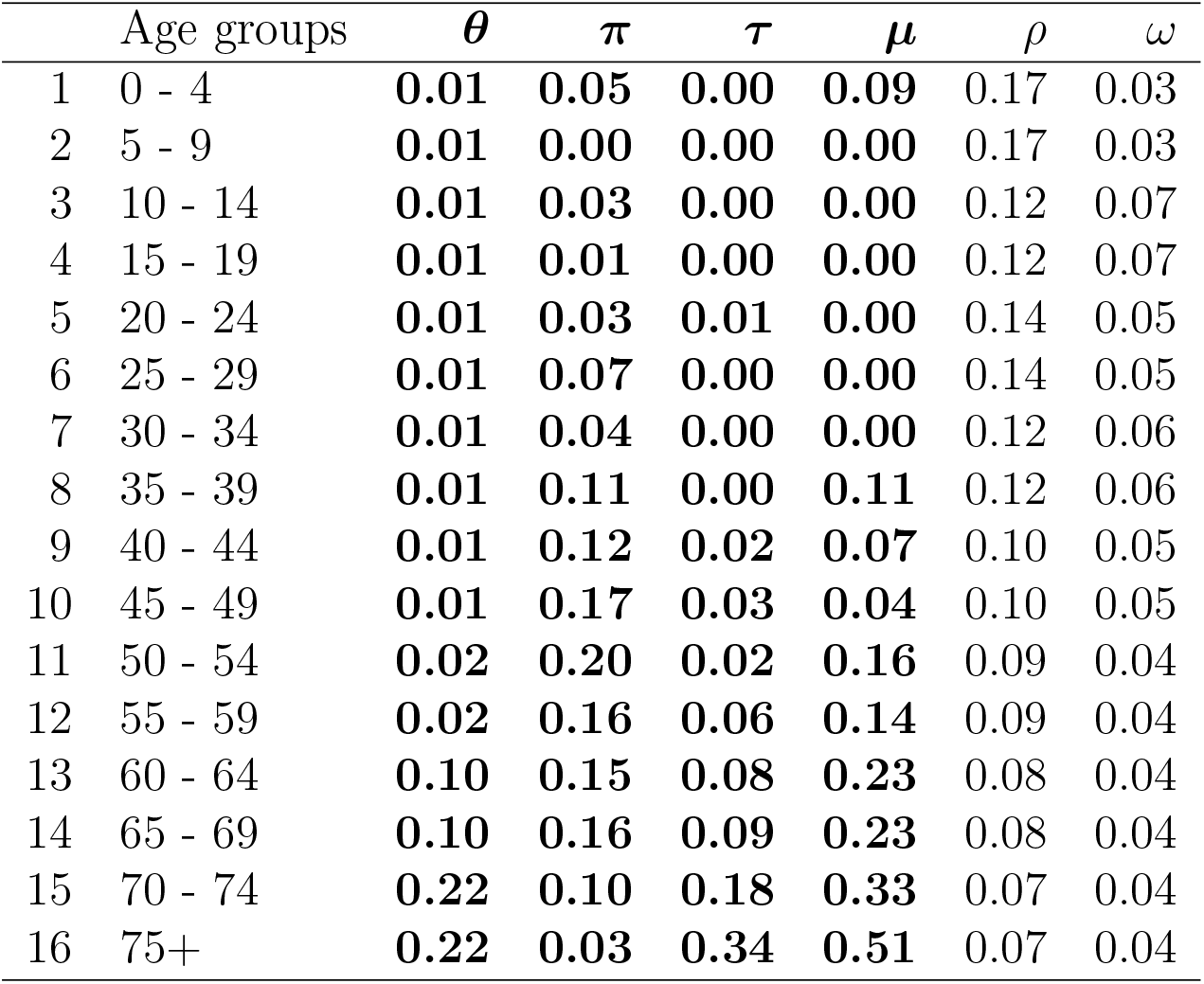
Age-group dependent parameters, both estimated (highlighted) and fixed for the 1-year immunity duration scenario.

| | Age groups | $\theta$ | $\pi$ | $\tau$ | $\mu$ | $\rho$ | $\omega$ |
| --- | --- | --- | --- | --- | --- | --- | --- |
| 1 | 0 - 4 | **0.01** | **0.05** | **0.00** | **0.09** | 0.17 | 0.03 |
| 2 | 5 - 9 | **0.01** | **0.00** | **0.00** | **0.00** | 0.17 | 0.03 |
| 3 | 10 - 14 | **0.01** | **0.03** | **0.00** | **0.00** | 0.12 | 0.07 |
| 4 | 15 - 19 | **0.01** | **0.01** | **0.00** | **0.00** | 0.12 | 0.07 |
| 5 | 20 - 24 | **0.01** | **0.03** | **0.01** | **0.00** | 0.14 | 0.05 |
| 6 | 25 - 29 | **0.01** | **0.07** | **0.00** | **0.00** | 0.14 | 0.05 |
| 7 | 30 - 34 | **0.01** | **0.04** | **0.00** | **0.00** | 0.12 | 0.06 |
| 8 | 35 - 39 | **0.01** | **0.11** | **0.00** | **0.11** | 0.12 | 0.06 |
| 9 | 40 - 44 | **0.01** | **0.12** | **0.02** | **0.07** | 0.10 | 0.05 |
| 10 | 45 - 49 | **0.01** | **0.17** | **0.03** | **0.04** | 0.10 | 0.05 |
| 11 | 50 - 54 | **0.02** | **0.20** | **0.02** | **0.16** | 0.09 | 0.04 |
| 12 | 55 - 59 | **0.02** | **0.16** | **0.06** | **0.14** | 0.09 | 0.04 |
| 13 | 60 - 64 | **0.10** | **0.15** | **0.08** | **0.23** | 0.08 | 0.04 |
| 14 | 65 - 69 | **0.10** | **0.16** | **0.09** | **0.23** | 0.08 | 0.04 |
| 15 | 70 - 74 | **0.22** | **0.10** | **0.18** | **0.33** | 0.07 | 0.04 |
| 16 | 75+ | **0.22** | **0.03** | **0.34** | **0.51** | 0.07 | 0.04 |

**Table 6:** Age-group dependent parameters, both estimated (highlighted) and fixed for the 3-year immunity duration scenario.

| | Age groups | $\theta$ | $\pi$ | $\tau$ | $\mu$ | $\rho$ | $\omega$ |
| --- | --- | --- | --- | --- | --- | --- | --- |
| 1 | 0 - 4 | **0.01** | **0.03** | **0.00** | **0.09** | 0.17 | 0.03 |
| 2 | 5 - 9 | **0.01** | **0.00** | **0.00** | **0.00** | 0.17 | 0.03 |
| 3 | 10 - 14 | **0.01** | **0.02** | **0.00** | **0.00** | 0.12 | 0.07 |
| 4 | 15 - 19 | **0.01** | **0.00** | **0.00** | **0.00** | 0.12 | 0.07 |
| 5 | 20 - 24 | **0.01** | **0.02** | **0.01** | **0.00** | 0.14 | 0.05 |
| 6 | 25 - 29 | **0.01** | **0.05** | **0.00** | **0.00** | 0.14 | 0.05 |
| 7 | 30 - 34 | **0.01** | **0.08** | **0.00** | **0.00** | 0.12 | 0.06 |
| 8 | 35 - 39 | **0.01** | **0.21** | **0.00** | **0.11** | 0.12 | 0.06 |
| 9 | 40 - 44 | **0.01** | **0.16** | **0.02** | **0.07** | 0.10 | 0.05 |
| 10 | 45 - 49 | **0.01** | **0.21** | **0.03** | **0.04** | 0.10 | 0.05 |
| 11 | 50 - 54 | **0.02** | **0.15** | **0.01** | **0.16** | 0.09 | 0.04 |
| 12 | 55 - 59 | **0.02** | **0.12** | **0.06** | **0.14** | 0.09 | 0.04 |
| 13 | 60 - 64 | **0.15** | **0.11** | **0.07** | **0.23** | 0.08 | 0.04 |
| 14 | 65 - 69 | **0.15** | **0.12** | **0.08** | **0.23** | 0.08 | 0.04 |
| 15 | 70 - 74 | **0.27** | **0.08** | **0.17** | **0.33** | 0.07 | 0.04 |
| 16 | 75+ | **0.27** | **0.03** | **0.32** | **0.51** | 0.07 | 0.04 |

The estimates for the time periods and respective change in population contacts, relative to pre-pandemic contacts, for each scenario calibration are presented in Table 7.

**Table 7:** Estimates for the periods and respective change in population contacts, relative to pre-pandemic contacts, for each calibration. S1, S2, S3 are in respect to the calibrations for the no-immunity-loss scenario, 1-year immunity duration scenario and 3-year immunity duration scenario.

| Scenario | $t_i$ | period | contact reduction ($l_i$) |
| --- | --- | --- | --- |
| S1 | 0-32 | 2021-01-24 - 2021-02-25 | 85.27% |
|  | 33-139 | 2021-02-26 - 2021-06-12 | 66.57% |
|  | 140-191 | 2021-06-13 - 2021-08-03 | 62.60% |
|  | 192-248 | 2021-08-04 - 2021-09-29 | 60.97% |
| S2 | 0-43 | 2021-01-24 - 2021-03-08 | 82.30% |
|  | 44-158 | 2021-03-09 - 2021-07-01 | 60.70% |
|  | 159-187 | 2021-07-02 - 2021-07-30 | 68.98% |
|  | 188-248 | 2021-07-31 - 2021-09-29 | 59.55% |
| S3 | 0-39 | 2021-01-24 - 2021-03-04 | 82.95% |
|  | 40-160 | 2021-03-05 - 2021-07-03 | 66.63% |
|  | 161-203 | 2021-07-04 - 2021-08-15 | 61.00% |
|  | 204-248 | 2021-08-16 - 2021-09-29 | 66.52% |

### Results discussion

Contacts were highly reduced during the first period due to the implementation of a nation-wide lockdown on January 15th and the closing of schools on January 22nd. Pre-schools opened on March 15th followed by the opening of schools on the 4th of April. Each calibration presents prior but close dates for the first change point, which might be explained by a reduced risk perception as cases went down and the phase-off of NPIs. For the no-immunity-loss scenario, we estimated the first change date as 2021-02-26, 1-year: 2021-03-09, 3-years: 2021-03-05. Estimated changes in contacts in the next period display an increase in contacts compared to the previous period. Change in contacts during this period was similar across all calibrations. The second change point occurs during June 13th, July 2nd and July 4th for the no-immunity-loss scenario 1-year immunity loss scenario and 3-year immunity loss scenario respectively. These dates occur after the 4th phase of lockdown lift on May 5th. The final change point occurred on August 4th (no-immunity loss scenario), July 31st (1-year immunity duration scenario) and August 16th (3-years immunity duration scenario). A small increase in contacts was observed in no-immunity-loss scenario and 1-year immunity duration scenario, which might result from few measures being in effect and the summer holidays. In the 3-year immunity scenario contact rates were at the same level as in period 2 (2021-03-05 to 2021-07-03) though we can see in figures 2 and 3 an inversion in the growing tendency. This of the effect of the vaccination campaign that reached a coverage 72% by the third week of October [15].

## Appendix C Simulation results

Figure 6 presents the evolution of the percentage of vaccine protected individuals in each age-group for each scenario. As expected, longer vaccine protection duration results in higher percentage of protected individuals as time goes on. These results also include the administration of a booster shot to those above 65 years of age. We can observe that by giving this dose we delay the decay of protection in this group for the 1-year and 3-year immunity duration scenarios.

**Figure 6:**
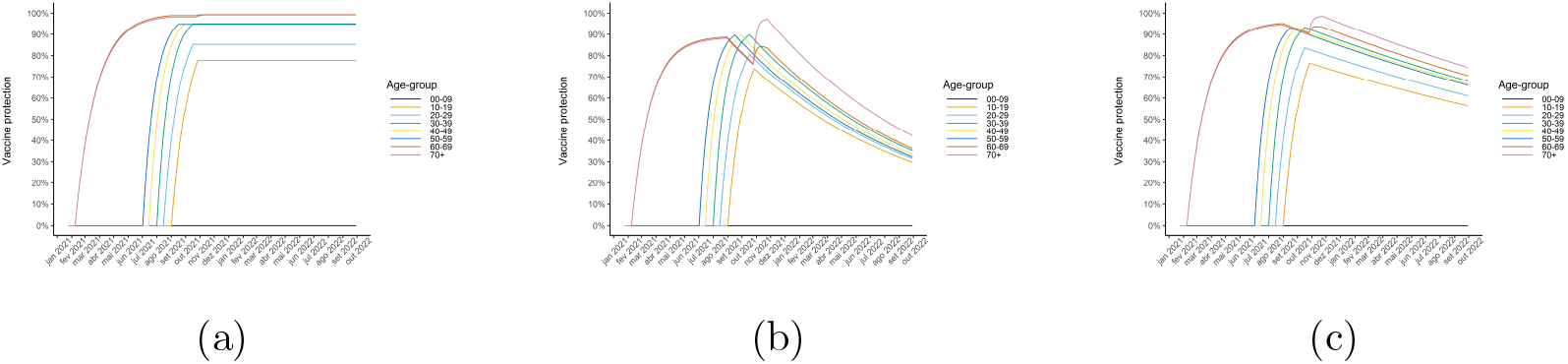
Percentage of vaccine protected individuals as a function of time, for each age-group for the no-immunity-loss (a), 1-year immunity duration (b) and 3-years immunity duration (c) scenarios with booster shot.

Figure 7 compares the evolution of ICU and non-ICU hospitalisations with and without a booster shot, by vaccine status, in the 1-year and 3-year immunity duration scenarios. This comparison encompasses a one year period between September 30th 2021 and September 30th 2022. We observe that administering the booster dose greatly reduces hospitalisations in the non-vaccinated groups while increasing the hospitalisations in the vaccinated group by a small amount.

**Figure 7:**
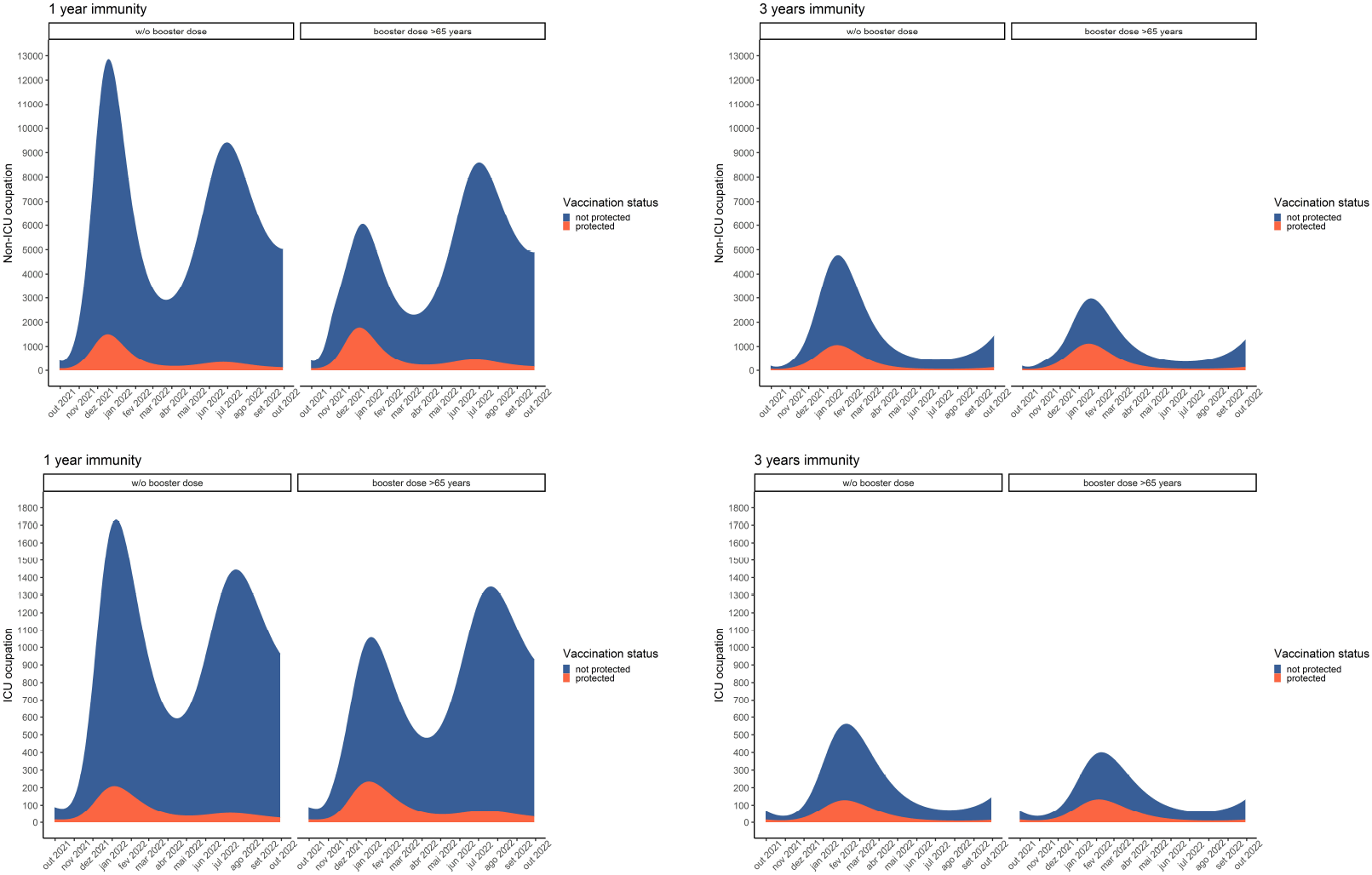
Evolution of the number of Non-ICU (top) and ICU (bottom) cases in the 1-year duration immunity scenario (left) and 3-year immunity duration scenario (right), with and without a third booster shot (*>* 65 years). Individuals are categorised as protected by vaccine and non-protected by the vaccine.

Figures 8 and 9 track the evolution of non-ICU and ICU cases by age-group during the period mentioned above, for the 1-year and 3-year immunity duration scenarios. We can observe that in the 3-year immunity duration scenario the prevalence of individuals above 70 years old in ICU and non-ICU care is reduced by administering a booster dose of the vaccine and stays below the scenario where no booster dose was administered. We can also observe the number of patients in these groups is reduced by administering the booster dose. In the 1-year immunity scenario we also observe that the administration of the booster dose reduces the prevalence of individuals above 70 years in non-ICU and ICU care, although by the end of the study period it reaches values similar to the scenario where no booster dose was administered. This is due to the faster loss of immunity (1-year duration) after receiving the booster dose.

**Figure 8:**
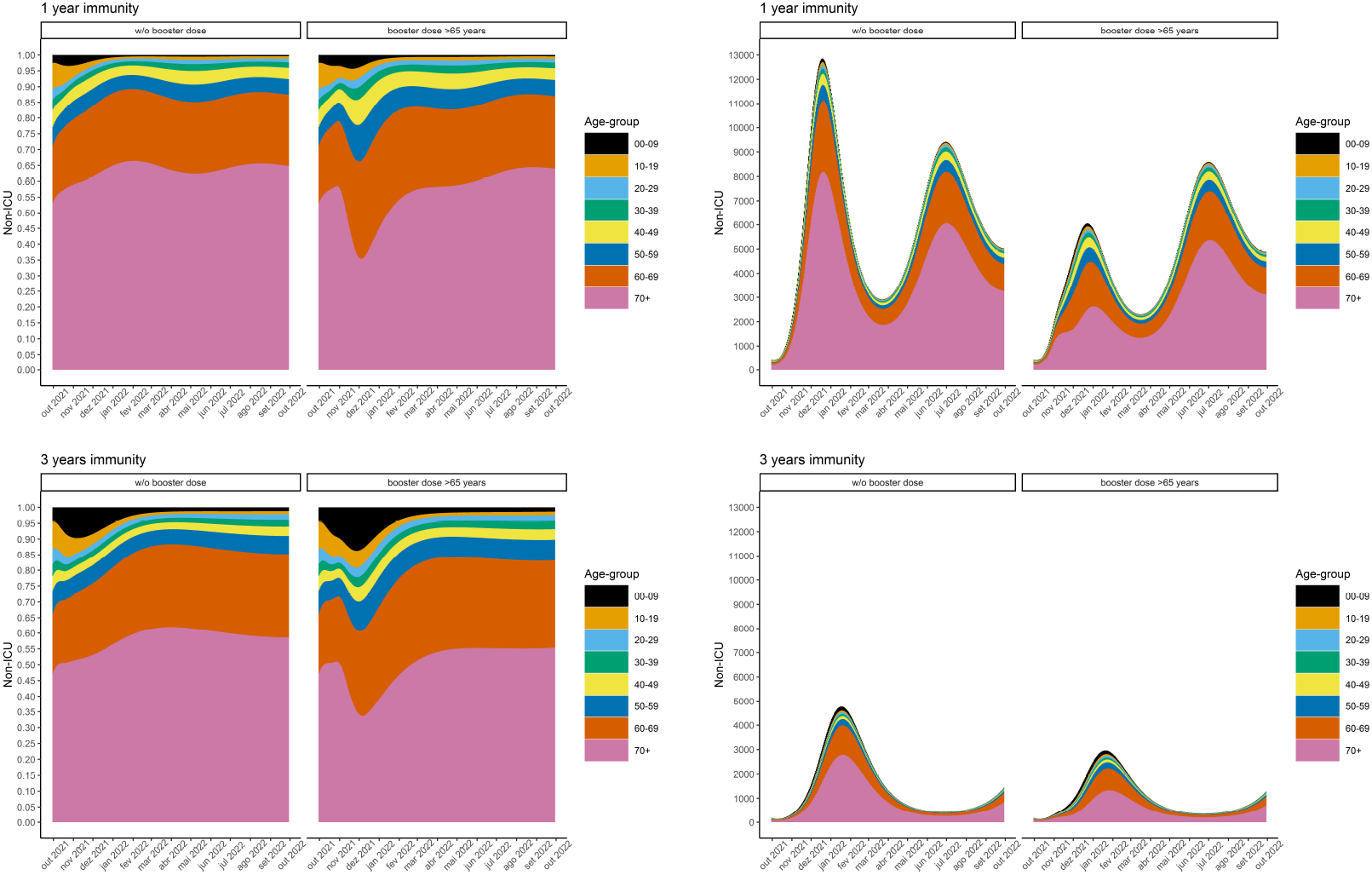
Evolution of the demography of non-ICU patients (left) and stacked number of non-ICU cases (right) by age-group for the 1-year immunity duration scenario (top) and 3 year immunity duration scenario (bottom).

**Figure 9:**
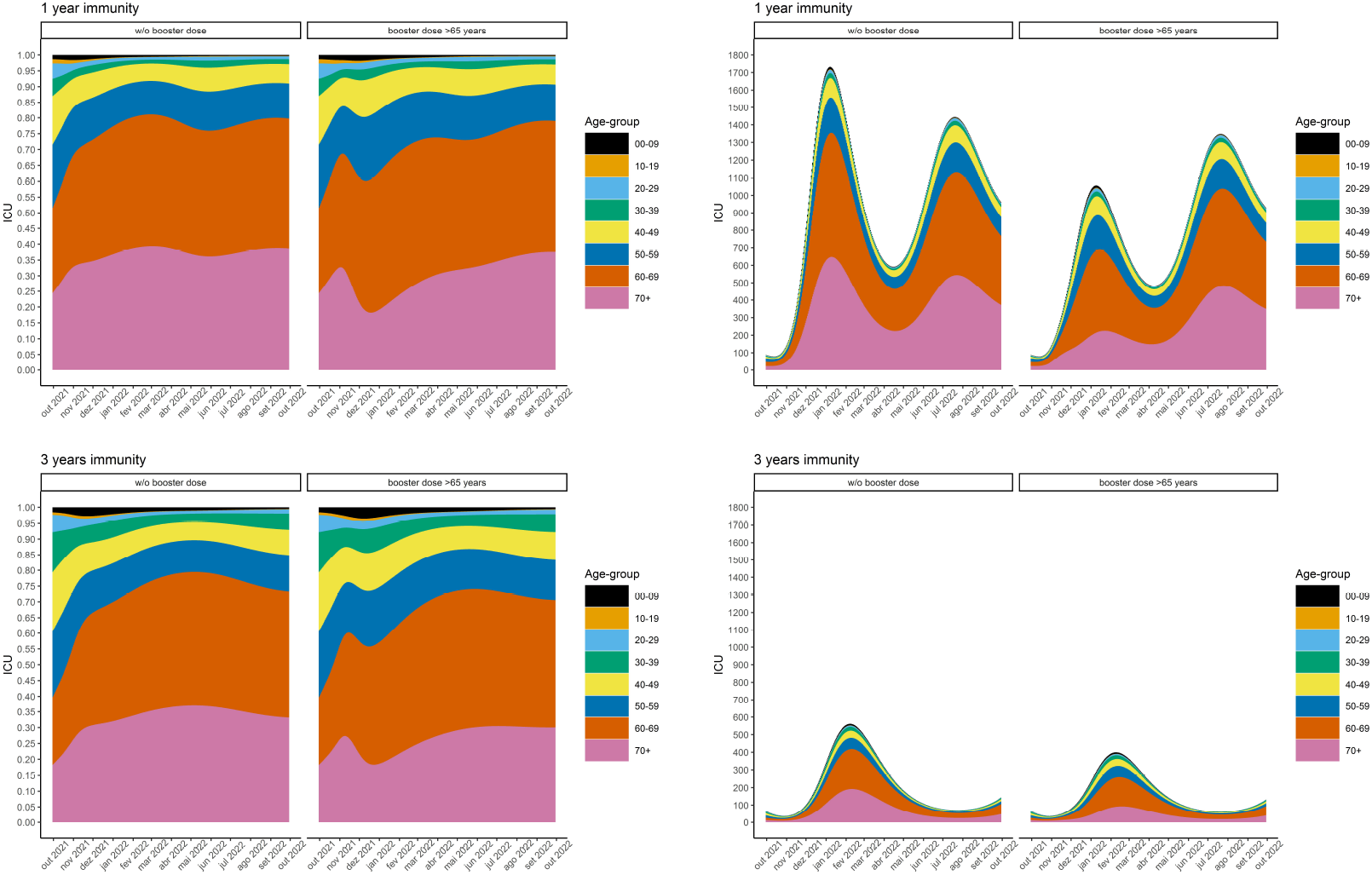
Evolution of the age demographics of ICU patients (left) and stacked number of ICU cases (right) by age-group for the 1-year immunity duration scenario (top) and 3 year immunity duration scenario (bottom).

## Funding

The authors acknowledge financial support from the Fundação para a Ciência e Tecnologia - FCT through project 692 2^a^ edição Research 4 covid, project name Projeção do Impacte das medidas Não-farmacológicas de Controlo e mitigação da epidemia de COVID-19 em Tempo ReaL (COVID-19 in-CTRL). The first author also acknowledges FCT within the PhD grants “DOCTOR-ATES 4 COVID”, number 2020.10172.BD. The second author also acknowledges FCT within projects UIDB/04621/2020 and UIDP/04621/2020. The third author also acknowledges FCT within the Strategic Project UIDB/00297 /2020 (Centro de Matemática e Aplicações, Universidade Nova de Lisboa).

## Notes

### Competing Interest Statement

The authors have declared no competing interest.

### Funding Statement

The authors acknowledge financial support from the Fundacao para a Ciencia e Tecnologia - FCT through project 692 2a edicao Research 4 covid, project name Projecao do Impacte das medidas Nao-farmacologicas de Controlo e mitigacao da epidemia de COVID-19 em Tempo ReaL (COVID-19 in-CTRL). The first author also acknowledges FCT within the PhD grants "DOCTORATES 4 COVID", number 2020.10172.BD. The second author also acknowledges FCT within projects UIDB/04621/2020 and UIDP/04621/2020. The third author also acknowledges FCT within the Strategic Project UIDB/00297/2020 (Centro de Matematica e Aplicacoes, Universidade Nova de Lisboa)

### Author Declarations

This study was approved by the Ethical Committee (Comissao de Etica para a Saude) of the Portuguese National Institute of Health.

